# Self-reported impulsivity, task-based inhibitory control, and early sipping behaviors as longitudinal predictors of adolescent alcohol use and problems in the ABCD Study

**DOI:** 10.64898/2026.02.03.26345434

**Authors:** Veronica Szpak, Everett L. Delfel, Alexander L. Wallace, Ryan M. Sullivan, Joanna Jacobus, Susan F. Tapert, Natasha E. Wade

**Affiliations:** SDSU / UC San Diego Joint Doctoral Program in Clinical Psychology; University of California, San Diego, Department of Psychiatry

**Keywords:** alcohol, problematic alcohol use, adolescents, impulsivity, inhibitory control

## Abstract

**Background:** Early low-level alcohol use predicts subsequent alcohol use and problems. Impulsivity and poor inhibitory control also predict later problematic alcohol use. However, few studies prospectively examine early sipping in combination with modeling impulsivity and inhibitory control change over time as predictors of adolescent alcohol use.

**Methods:** Data Release 6.0 from the Adolescent Brain Cognitive Development (ABCD) Study was used (n=11,866; 48% Female). A series of linear mixed-effect models examined trajectories of non-religious sipping at baseline (ages 9-10) and self-reported impulsivity (UPPS-P) and task-based inhibitory control (Flanker task) over time as predictors of past year drinks and problematic alcohol use by ages 15-16. Predictors were run as separate models and a full model with all predictors together. Models were nested within the participant and study site. Interactions with age (to measure change over time from Baseline to Year 6) were included. Corrections for multiple comparisons were employed.

**Results:** In individual models, four impulsivity interactions were significant: (1) negative urgency*age (β=.04, FDR-*p*<.001), (2) positive urgency*age (β=.04, FDR-*p*<.001), (3) lack of planning*age (β=.04, FDR-*p*<.001), and (4) sensation seeking*age (β=.04, FDR-*p*<.001), suggesting that as age increases, the relationship between impulsivity and alcohol use strengthens. Sipping*age (β=.02, FDR-*p*<.001) interactions also predicted standard drinks. Regarding problematic use, there was a significant interaction in the full model: negative urgency*age (β=-.07, *p*=.05), indicating that this relationship is more pronounced at earlier ages.

**Conclusions:** Trait impulsivity and sipping in late childhood relate to future alcohol use, and the relationship strengthens with age. Our results found a negative interaction between negative urgency and age on problematic use, potentially indicating negative urgency as a phenotype of vulnerability to experiencing alcohol related problems at younger ages. Findings indicate the importance of understanding facets of impulsivity in the context of adolescent alcohol use for prevention and intervention efforts.

## Introduction

In 2023, the National Survey of Drug Use and Health reported that 12.6 million (33.1%) youth between the ages of 12 and 20 have had at least one drink in their lives (SAMHSA). Several risk factors are associated with increasing alcohol use and problems across adolescence: drinking alcohol at an early age, even when only sipping (Donovan & Molina, 2011), impulsivity, and inhibitory control (Quach et al., 2020). Yet these three factors are limitedly considered together across developmental stages. A multi-method approach that includes behavioral (early, low-level alcohol use), trait-based self-report (impulsivity), and task-based (inhibitory control) predictors may clarify the relative utility of each factor in predicting alcohol use and related problems over time. One study that utilizes this multi-method approach is the Adolescent Brain Cognitive Development Study (ABCD) a longitudinal study that follows that same cohort over 10 years (Lisdahl et al., 2018). Therefore, ABCD provides a valuable opportunity to study these behavioral, trait-based, and task-based predictors.

Research shows that sipping behavior in youth predicts consuming full drinks in later adolescence (Donovan & Molina, 2011; Jackson et al., 2015). For instance, Donovan and Molina (2011) found that sipping alcohol by the age of 10 predicted early-onset drinking, defined as age 14 or younger, compared to non-sippers. Similarly, Jackson and colleagues (2015) found that youth who sipped alcohol in the sixth grade had an increased risk of having a full drink, getting drunk, and drinking heavily by the ninth grade. Consistent with this, studies within ABCD show that sipping at earlier ages and traits such as impulsivity predict alcohol use in adolescence (Choi et al., 2025; May et al., 2022). Given that there is a developmental progression in alcohol use, understanding the potential consequences of early alcohol initiation is crucial (Donovan & Molina, 2011). However, it is unclear how sipping or low-level alcohol use in early adolescence, alongside other risk factors, predicts problematic use later in adolescence.

Trait impulsivity is another risk factor for alcohol use in adolescents. A common measure is the Urgency Premeditation Perseverance Sensation Seeking (UPPS-P) scale, which consist of five subscales (positive urgency, negative urgency, sensation seeking, lack of perseverance, and lack of planning) to delineate separate facets of impulsivity. A series of studies using the UPPS-P found that all subscales are associated with early alcohol sipping by ages 9-10 (Watts et al., 2021) and sensation seeking is associated with problematic alcohol use across adolescence (Fernández-Artamendi et al., 2018; Niklason et al., 2025; Watts et al., 2024). Additional studies focusing on college-age students found that positive and negative urgency are also linked to different domains of alcohol problems (Jones et al., 2014; McCarty et al., 2017; Tran et al., 2018). This may be due to adolescents using alcohol to cope with negative emotions or to enhance positive emotions as they transition into adulthood. Further research on impulsivity and alcohol use patterns in adolescents is needed to understand these trajectories.

Additionally, inhibitory control, often measured through neurocognitive tasks, is another risk factor for problematic alcohol use in adolescents. A longitudinal study found that poor response inhibition on the Stopping Task at ages 12-14 predicted the onset of alcohol use-related problems in adolescence at ages 15-17 (Nigg et al., 2006). Adolescence is considered a vulnerable stage in life due to ongoing neurodevelopment, which may impact inhibitory control, leading to difficulties in regulating alcohol use. Conversely, a narrative review of a series of studies suggests that there is a bidirectional relationship between inhibitory control and alcohol-related problems, such that alcohol use may impact inhibitory control performance or that poor inhibitory control may predict alcohol use-related problems (López-Caneda et al., 2014). Longer trajectories of inhibitory control and alcohol use are needed to refine understanding of these relationships.

Few studies have assessed whether presence of lifetime alcohol sipping across childhood or evolving impulsivity and inhibitory control facets across adolescence predict adolescent alcohol use or related problems into later adolescence. In addition, there is a lack of studies examining the relationship among these three risk factors in tandem. Therefore, this study aims to use the ABCD study to accomplish the following objectives: (1) to examine whether early sipping, impulsivity, and inhibitory control predict trajectories of increased alcohol use through year 6 follow-up (ages 15-16), and (2) to examine if these same factors predict problematic alcohol use through year 6. We expect that early sipping, impulsivity, and inhibitory control will be significant predictors of increased and problematic alcohol use. In addition, we expect that higher impulsivity scores and poorer inhibitory control performance will be associated with increased and problematic alcohol use.

## Methods and Materials

### Dataset

11,880 youth (ages 9-10 at baseline) were enrolled in the ABCD study, which is a 10-year multi-site study in the United States examining adolescent brain and cognitive development across the U.S. Data collection includes cognitive, social, emotional, environmental, behavioral, and academic assessments (Volkow et al., 2018) as well as neuroimaging and biospecimens for hormonal, genetic, environmental exposure, and substance use analysis (Uban et al., 2018). Current analyses use the ABCD Data Release 6.0 (Jernigan et al., 2025).

### Study Session Procedure

Youth participants and their parent/guardian complete yearly follow-up visits (through year 6; ages 11-17). Study sessions include a series of questionnaires and neurocognitive tasks, biospecimen collection, and a biannual MRI scan (Barch et al., 2018; Lisdahl et al., 2018; Luciana et al., 2018; Uban et al., 2018). Study sessions are completed both in-person, fully remote, and hybrid (e.g., remote interviews/questionnaires, in-person bioassays and neuroimaging collection).

### Materials

#### Demographics

Sex assigned at birth, race/ethnicity, and parental education were collected at baseline. Age was collected at each annual assessment (Barch et al., 2018).

#### NIH Toolbox Flanker Task

Participants used an iPad with the assistance of trained research staff to complete the NIH Toolbox Flanker Task (Beaumont et al., 2013). This task measures sustained attention and inhibitory control. The participant is presented with a target stimulus in the center of a row of stimuli and instructed to select the direction of the target stimulus while ignoring the other stimuli (Luciana et al., 2018). An age-corrected standard score of combined accuracy and reaction time was used as a primary predictor in models.

#### Urgency-Premeditation-Perseverance-Sensation Seeking (UPPS-P)

The UPPS-P examines five facets of impulsivity (4 items per facet) by youth self-report: negative urgency, positive urgency, lack of planning, sensation seeking, and lack of perseverance. Participants complete the short form of the UPPS-P every two years, beginning at baseline (ages 9-10). The response format ranges from 1 (*not at all like me*) to 4 (*very much like me*) (Barch et al., 2018). Higher scores indicate stronger impulsivity. Total score on each subscale was used in analytical models.

#### Substance Use Interview

During the baseline visit, participants who had heard of alcohol were asked, “*Have you had a sip of alcohol such as beer, wine, or liquor (rum, vodka, gin, whiskey)”* (in their lifetime) from the iSay Sip Inventory. When endorsed, a follow-up question queried whether they ever sipped outside of a religious ceremony. Participants who endorsed sipping were then asked, “*Have you had a full drink of alcohol such as beer, wine, or liquor (rum, vodka, gin, whiskey)”.* Consistent with prior research (Watts et al., 2021), only non-religious sipping is included here. At annual follow-up visits, the same questions were queried for use since their last visit.

#### Timeline Followback (TLFB)

A web-based version of the Timeline Followback is administered by trained research staff to assess substance use patterns (e.g., product, dose, and routes of administration) (Lisdahl et al. 2018). The past 6 months are assessed at baseline, and time since last study visit are assessed during follow up sessions. Participants who endorsed standard drinks of alcohol were prompted to complete the Timeline Followback to record standard drinks of use since their last visit, as well as additional measures regarding attitudes and consequences of alcohol use (Lisdahl et al., 2018). Here, the number of standard alcohol drinks since their last visit (after baseline) were used as a primary outcome variable. Cannabis and nicotine use since their last visit were included in secondary models as covariates.

#### Rutgers Alcohol Problem Index (RAPI)

All participants endorsing alcohol use completed the RAPI to examine the frequency of negative consequences related to adolescent drinking in the past year. The RAPI has 23 items and uses a Likert scale ranging from 0 (*never*) to 4 (*more than 10 times*) (Lisdahl et al., 2018; White & Labouvie, 1989). Higher scores indicate more alcohol-related negative consequences. The total sum score of the RAPI was used as an outcome variable.

### Data Analysis Plan

R and RStudio were used for all analyses (RStudio Team, 2025). Decisions on statistical significance were set at p≤.05.

#### Data Cleaning and Multiple Imputations

Multiple imputations were performed for impulsivity (2.28% missingness), inhibitory control (15.59% missingness), and parental education (0.1% missingness) using the *mice* package (Buuren & Groothuis-Oudshoorn, 2011) in R.

#### Primary Analyses of Past Year Drinking Patterns

All models were constrained to baseline and even years of visits (i.e., years 2, 4, and 6) given that the UPPS-P and the Flanker task were only administered at these visits. Standardized beta coefficients were calculated in R using the *effectsize* package in R (Ben-Shachar et al., 2020). A series of linear mixed-effects models separately examined trajectories of non-religious sipping at baseline (ever sipped in lifetime, yes/no; ages 9-10), impulsivity (each UPPS-P subscale independently), and inhibitory control (Flanker) as predictors of total number of past year drinks (TLFB). A full mixed effects model was also conducted combining all predictor variables (i.e., sipping, impulsivity subscales, and inhibitory control) to predict total number of past-year alcohol drinks. Interaction of primary predictors with age (to account for time from baseline to year 6) were included. Models were nested within the participant and study site. Age and sex assigned at birth are included as covariates for all individual models. Due to convergence issues, the full model did not include sex. Though considered, past year other substance use (nicotine and cannabis) was not included in the primary models due to convergence issues; however, coefficients on non-converging models were similar to primary reported models. False Discovery Rate (FDR) corrections accounted for multiple comparisons (i.e., 8 primary models) using the Benjamini-Hochberg method (Wright, 1992).

#### Secondary Analyses of Problematic Alcohol Use

Secondary models are limited to those with significant predictors from the primary separate models. The same modeling structure with nesting and covariates were used. Past year substance use (nicotine and cannabis) was added as additional covariates. Linear mixed-effect models assess whether each factor independently predicts RAPI score. Further, a full model with all significant predictors (including interaction with age for each individual factor) was run to assess which factors predict RAPI score in the context of accounting for all additional variables.

## Results

### Participant Characteristics

The total sample included in analyses consisted of 11,866 participants. Nearly half (48%) were female. **Table 1** describes additional demographic and substance use information. Participants who reported at least one full standard drink were more likely to be female (53%; *p* < .001) and white (78%; p < .001). Descriptive statistics for lifetime total alcohol drinks within those who drink were as follows: M = 6, SD = 51.6, range: 1-2,552. The mean RAPI score was 2 (SD = 3.5, range: 0-31). See **Table 2** for group differences between participants who drank alcohol and those who never drank.

**Table 1.** Full sample and RAPI sample participant characteristics.

| *Participant Characteristics* | Full Sample<br>N = 11,866 | RAPI Sample<br>N = 660 |
| --- | --- | --- |
| Sex, Female, *n*, (%) | 5,678 (47.8%) | 371 (56.2%) |
| Race, *n*, (%) |  |  |
| White | 7,805 (65.7%) | 543 (82.3%) |
| Black | 1,929 (16.3%) | 24(3.6%) |
| Asian | 256 (2.2%) | <10 (<5%) |
| AI/AN & NHPI (Combined) | 78 (0.6%) | < 10(<5%) |
| More than One Race | 1,280 (10.8%) | 64 (9.7%) |
| Other | 518 (4.4%) | 20 (3%) |
| Ethnicity, *n*, (%) |  |  |
| Hispanic | 2,445 (20.6%) | 107 (16.2%) |
| Non-Hispanic | 9,379 (79%) | 552 (83.6%) |
| Missing | 42 (0.4%) | <10 (<5%) |
| Parental Education, *n*, (%) |  |  |
| 12<sup>th</sup> Grade and Below | 593 (5%) | <10 (<5%) |
| High School Graduate/ GED or Equivalent | 1,132 (9.5%) | 35 (5.3%) |
| Associate Degree/ Some College | 3,074 (25.9%) | 138 (20.9%) |
| Bachelor's Degree | 3,013 (25.4%) | 182 (27%) |
| Graduate School (e.g., MA, MD, PhD) | 4,042 (34.1%) | 296 (44.8%) |
| Missing | <20 (<5%) |  |

**Table 2.** Group differences in those who drank relative to those who did not drink.

| **Variable** | **No Alcohol Use**<br>**N = 10,429<sup>1</sup>** | **Alcohol Use**<br>**N = 1,439<sup>1</sup>** | **p-value<sup>2</sup>** |
| --- | --- | --- | --- |
| **Sex** |  |  | <0.001 |
| Male | 5,512 (53%) | 678 (47%) |  |
| Female | 4,917 (47%) | 761 (53%) |  |
| **Race** |  |  | <0.001 |
| White | 6,687 (64%) | 1,118 (78%) |  |
| Black | 1,839 (18%) | 90 (6.3%) |  |
| Asian | 244 (2.3%) | <20 (<5%) |  |
| AI/AN & NHPI (Combined) | 67 (0.6%) | <20 (<5 %) |  |
| More than One Race | 1,125 (11%) | 155 (11%) |  |
| Other | 465 (4.5%) | 53 (3.7%) |  |
| Missing | <10 (<5%) |  |  |
| **Ethnicity** |  |  | 0.4 |
| Hispanic | 2,158 (21%) | 287 (20%) |  |
| Non-Hispanic | 8,229 (79%) | 1,150 (80%) |  |
| Missing | 42 (0.4%) | <10 (<5%) |  |
| **Parental Education** |  |  | <0.001 |
| 12<sup>th</sup> Grade and Below | 559 (5.4%) | 34 (2.4%) |  |
| High School Graduate/ GED or Equivalent | 1,041 (10%) | 91 (6.3%) |  |
| Associate Degree/ Some College | 2,720 (26.1%) | 354 (24.6%) |  |
| Bachelor's Degree | 2,658 (25.5%) | 355 (24.6%) |  |
| Graduate School (MA, MD, PhD) | 3,437 (33%) | 605 (42%) |  |

*Note.* <sup>1</sup> n (%), <sup>2</sup> Pearson's Chi-squared test. Alcohol use is defined as reporting at least one standard drink.

### Primary Analyses of Past Year Alcohol Use Patterns

See **Figure 1** for all significant interactions in primary models.

**Figure 1.**
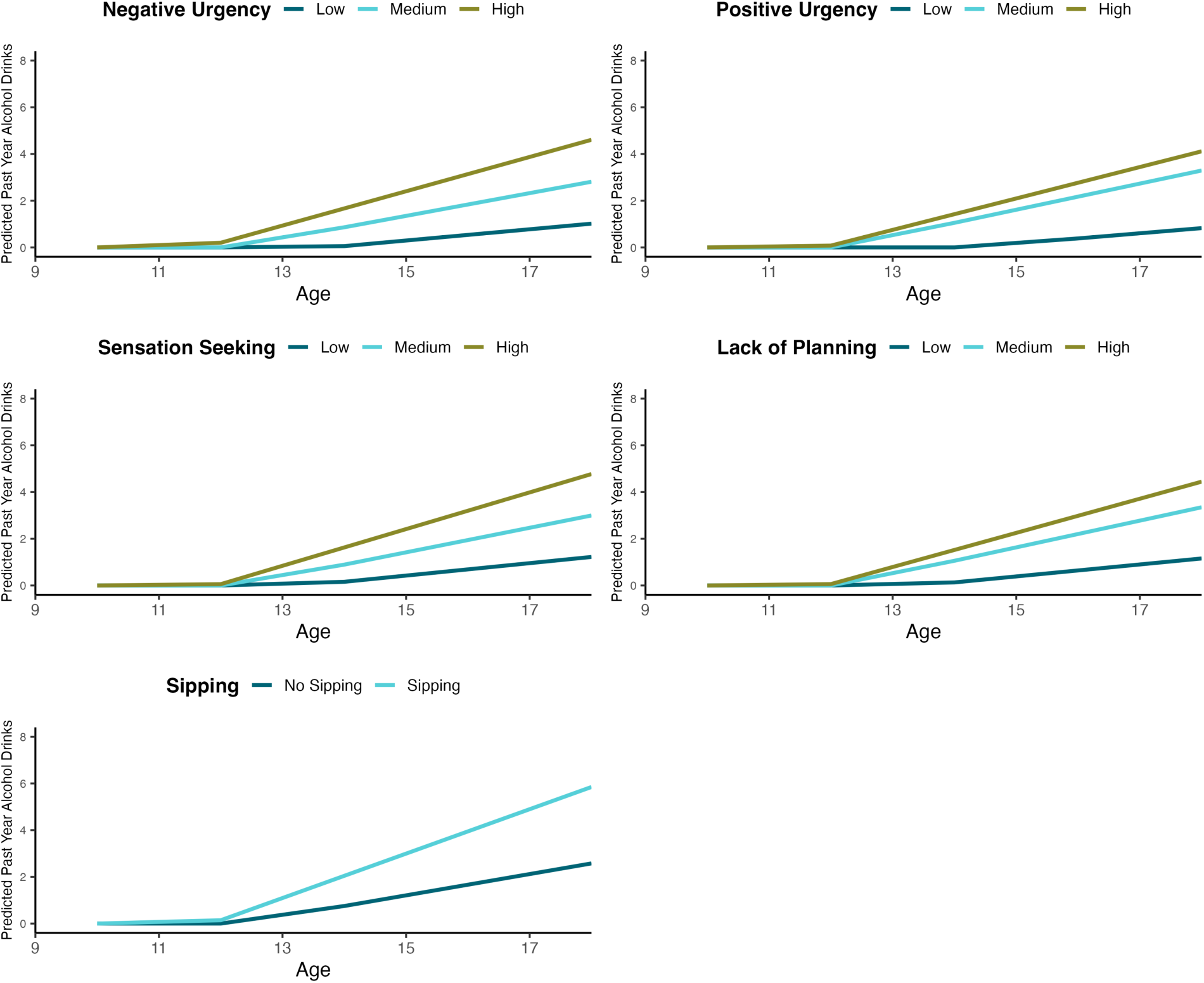
Interactions between age and each significant impulsivity subscale from primary individual models. ***Note.*** Predicted values from the fitted models are shown across visit age for three levels of each construct: UPPS-P subscales, corresponding to the 25th (Low), 50th (Medium), and 75th (High) percentiles.

#### Sipping at Baseline

The main effects of non-religious sipping (β = .01 SE = 1.44, *t* = - 3.92, *p* < .001, FDR *p < .*001) and age (β = .07, SE = .05, *t* = 9.59, *p* < .001, FDR *p <.* 001) were significant at predicting increased alcohol use since last visit. The interaction between sipping*age was also significant (β = .02, SE = .11, *t* = 4.40, *p* < .001, FDR *p < .*001), indicating that early sipping predicted increased alcohol use between visits across time.

#### UPPS-P

##### Negative Urgency

The main effects of negative urgency (β = .03, SE = .21, *t* = -6.28, *p <* .001, FDR *p* < .001) and age (β = .06, SE = .15, *t* = -3.37, *p* = <.001, FDR *p* = .001) were significant. The interaction between negative urgency*age was also significant (β = .04, SE = .02, *t* = 7.34 *p* <.001, FDR *p* < .001), such that higher negative urgency scores and older age predicted more drinks since last visit.

##### Positive Urgency

The main effects of positive urgency (β = .03, SE = .19, *t* = -6.38 *p* < .001, FDR *p* < .001) and age (β = .06, SE = .13, *t* = -2.70 *p* <.01, FDR *p < .*01) were significant. In addition, the interaction between positive urgency*age was significant (β = .04, SE = .02, *t* = 7.45, *p* < .001, FDR *p* < .001), such that higher positive urgency scores and older age predicted more drinks over time.

##### Lack of Perseverance

There was a significant main effect of age in predicting drinks since last visit (β = .07, SE = .14, *t* = 3.74 *p* < .001, FDR *p =.* 002). There was no significant main effect (*p* = .93) or interaction between lack of perseverance*age (*p* = .90).

##### Lack of Planning

The main effects of lack of planning (β = .03, SE = .24, *t* = -7.38, *p* < .001, FDR *p* < .001) and age (β = .07, SE = .15, *t* = -4.49, *p* < .001, FDR *p* < .001) were significant. In addition, the interaction between lack of planning*age was significant (β = .04, SE = .02, *t* = 8.32, *p* < .001, FDR *p* < .001), such that higher lack of planning urgency scores and older age predicted more drinks since last visit.

##### Sensation Seeking

The main effects of sensation seeking (β = .02, SE = .19, *t* = -7.28, p < .001, FDR *p* < .001), and age (β = .06, SE = .16, *t* = - 4.75, *p* < .001, FDR *p* < .001), were significant. In addition, the interaction between sensation seeking*age was significant, (β = .04, SE = .02, *t* = 8.29, *p* < .001, FDR *p* < .001), such that higher sensation scores and older age predicted more drinks since last visit.

#### Inhibitory Control

The main effect of inhibitory control performance was not significant in predicting alcohol drinks (*p* = .08). The interaction between inhibitory control*age also was not significant (*p* = .06).

#### Full Model

The main effects of sipping (β = .008, SE = 1.44, *t* = -3.07, *p* < .01, FDR *p* < .01), sensation seeking (β = .01, SE = .21, *t* = -4.63, *p* < .001, FDR *p* < .01), lack of perseverance (β = -.01, SE = 0.27, *t* = 3.19, *p* < .01, FDR *p* = .03), lack of planning (β = .02, SE = .28, *t* = -5.84, *p* < .001, FDR *p* < .001), negative urgency (β = .02, SE = .25, *t* = -3.01, *p* < .01, FDR *p* = .003) and positive urgency (β = .01, SE = .23, *t* = -1.98, *p* = .047, FDR *p* = .047) were significant in predicting alcohol drinks since last visit. There was also a significant main effect of age (β = .06, SE = 0.37, *t* = -5.85, *p* < .01, FDR *p* < .01). Inhibitory control performance (*p* = .27) was not significant.

The interaction between baseline sipping*age was significant (β = .02, SE = .11, *t* = 3.43, *p* < .001, FDR *p* < .01) in predicting alcohol drinks since last year. Multiple UPPS-P and age interactions were significant: sensation seeking*age (β = .03, SE = .02, *t* = 5.11, *p* < .001, FDR *p* < .001), lack of perseverance*age (β = - .01, SE = .02, *t* = -3.68, *p* < .001, FDR *p* = .01), lack of planning*age (β = .03, SE = .02, *t* = 6.47, *p* < .001, FDR *p* < .001), positive urgency*age (β = .01, SE = .02, *t* = 2.33, *p* = .02, FDR *p* = .02), negative urgency*age (β = .03, SE = .02, *t* = 3.52, *p* < .001, FDR *p* < .001). Each showed the same pattern as in the individual models, except for lack of perseverance, which demonstrated an association in the opposite direction. Inhibitory control*age was not significant (*p* = .25).

### Secondary Analysis of Problematic Alcohol Use

#### Independent RAPI Models

There were no significant main effects of the UPPS-P subscales predicting total RAPI score: negative urgency (*p* = .11), positive urgency (*p* = .51), lack of planning (*p* = .85), and sensation seeking (*p* = .44). There were also no significant interaction effects with age: Negative urgency*age (*p* = .22), positive urgency*age (*p* =.33), lack of planning*age (*p* = .97), and sensation seeking*age (*p* = .47). There was no significant main effect of sipping (*p* = .63) or its interaction with age (*p* = .66). Nicotine and cannabis use were significant in predicting RAPI score across all models (*p*s < .05).

#### Combined RAPI Model

In the full model with all the subscales and their interaction with age, there was a significant main effect of negative urgency (β =.14, SE = .84, *t* = 2.19, *p* = .02). There were no other main effects. There was also significant interaction effect between negative urgency*age (β = -.07, SE = .05, *t* = -1.96, *p* = .05) such that as adolescents age, negative urgency decreases. There were significant main effects of nicotine and cannabis use since last visit (*p* < .05).

## Discussion

This study had two aims: first, to examine whether measurements of early sipping behavior, self-reported trait impulsivity, and task-based inhibitory control predicted trajectories of increased alcohol use by ages 15-16; second, to examine if these same factors predict problematic alcohol use by ages 15-16. We hypothesized that sipping, impulsivity, and inhibitory control would predict increased and problematic alcohol use over time. Consistent with this, we found alcohol sipping, nearly all impulsivity subscales, and inhibitory control interacted with age in predicting alcohol use, with higher scores and older age predicting more past-year alcohol drinks. However, findings regarding problematic alcohol use were limited: there were no significant predictor and age interactions in separate models. When considering all risk factors together, negative urgency interacted with age in predicting problematic alcohol use.

Findings related to early sipping behavior were consistent with prior literature on predicting increased alcohol use in later adolescence (Donovan & Molina, 2011; Jackson et al., 2015). Across models, we found significant interactions of early non-religious sipping and age, as well as impulsivity and age. Youth that sip outside of family environments and score high on trait impulsivity may be more likely to consume full drinks earlier. This is consistent with the findings by Watts and colleagues (2024), which found that sipping was associated with impulsivity in the same cohort (i.e., ABCD). In regard to the RAPI, we did not find any significant effects related to problematic use, which contrasts with some of the early sipping research (Jackson et al., 2015). This may suggest that sipping alone may not be a risk factor of problematic use, but rather a sign of other underlying risk factors (e.g., impulsivity contributing to both sipping behavior and later use). Greater investigation into the contextual factors which moderate early sipping behavior and later and problematic alcohol use may clarify these relationships.

All impulsivity subscales (except for lack of perseverance) and their interaction with age were significant in the individual models predicting alcohol use, indicating that the relationship between increased alcohol use and impulsivity is conditional on age. As youth age and in those whose trait impulsivity increases, their alcohol use tends to increase. Our results also highlight that the relationship between impulsivity and alcohol use gets stronger over time. This is consistent with research that shows that high impulsivity scores on traits such as sensation seeking and positive and negative urgency are associated with higher levels of alcohol use during emerging adulthood (Shin et al., 2012), and with a meta-analysis that found all UPPS-P traits were associated with alcohol consumption in adolescents (Stautz & Cooper, 2013). This suggests that impulsivity tends to peak during adolescence, particularly between the ages of 12-4, which may be linked to alcohol use initiation. Additionally, while not explored here, the relationship between alcohol use and impulsivity could be bidirectional, with alcohol use predicting impulsivity (Kaiser et al., 2016). Future research should explore longitudinal directional approaches such as trajectory growth models or cross lagged panel models (Freichel et al., 2023).

Our results did not find an association between increased alcohol use and inhibitory control. This is inconsistent with other studies that have found that poorer inhibitory control tends to be associated with increased alcohol use (López-Caneda et al., 2014). It may be that more frequent and higher quantities of alcohol use impacts inhibitory control (López-Caneda et al., 2014), suggesting that the alcohol use reported in this cohort is too low to impact neurocognitive trajectories yet. Further, other research indicates that in those with an alcohol use disorder, developmental trajectories of inhibitory control were altered (Wilson et al., 2021), supporting the interpretation that the ABCD cohort is not using higher quantities of substance use for the relationship to be more evident. Taken together, the association between inhibitory control and alcohol use should be explored further in longitudinal studies with more regular alcohol use and in clinical samples.

In regard to problematic alcohol use, our results considering individual risk factors in predicting RAPI score revealed that negative urgency is associated with problematic alcohol use and interacts with age. Negative urgency refers to acting impulsively when experiencing distress or negative emotions. These findings may indicate that adolescents with problematic drinking behaviors may drink alcohol to relieve negative moods or emotional states, which is consistent with prior studies (VanderVeen et al., 2016). The neurocircuitry of addiction model suggests that negative urgency contributes to addiction through negative reinforcement (Zorrilla & Koob, 2019). Interestingly, however, findings here show that this relationship is stronger at younger ages. Prior research indicates that changes in impulsivity may occur as adolescents age (Quinn & Harden, 2013). Given this was only present in full models of all predictors, perhaps the relationship between negative urgency and alcohol problems is only evident once accounting for all other facets of impulsivity. Alternatively, negative urgency may capture a unique aspect of impulsivity that is not adequately represented by the other subscales. In either case, those most vulnerable to developing alcohol use problems may be those with high negative urgency at younger ages.

A strength of this analysis is that these risk factors are measured together in the same cohort, and most participants are substance naive during baseline (ages 9-10), allowing for assessment of changes in predictors and within participants as they age (Lisdahl et al., 2021). Results suggest a possible link between sipping, impulsivity, and inhibitory control on alcohol use across adolescence. Future directions for this line of research could examine how impulsivity is related to environment, peer substance use, family history, and attitudes toward substance use to get a more comprehensive understanding of what plays a role in early substance use initiation.

### Limitations

Sample size was restricted only to those who use alcohol for the secondary models; therefore, only 660 participants were prompted to complete the RAPI, consistent with expectations that using full standard drinks of alcohol is required to experience alcohol-related problems. Alcohol use is still also limited compared to later developmental stages (e.g., young adulthood) and the mean score on the RAPI for this sample is 2 (range: 0-31). Accordingly, even if participants were prompted to complete the RAPI, they may have not endorsed many items related to problematic alcohol use. Continued monitoring of the development of problematic alcohol and other substance use is needed. Despite the strengths of the multi-modal predictors, the outcome was limited to self-reported substance use, which may be under-reported in the ABCD cohort (Wade et al., 2023). While within-subject trajectories were measured here, directionality and causality cannot be determined; future models incorporating causal inference will provide further clarity on these relationships. Consistent with expectations for studies using the ABCD dataset, effect sizes were small (Dick et al., 2021).

## Conclusions

Overall, results indicate that the relationship between early non-religious alcohol sipping and certain subscales of impulsivity, with alcohol use vary across adolescent development. When considering alcohol related problems, negative urgency, a facet of impulsivity, is associated with problematic alcohol use with those with higher negative urgency at younger ages at risk of worse problematic alcohol use trajectories. Clinical assessment for problematic alcohol use should be implemented earlier to identify adolescents who are drinking alcohol at younger ages (12-13). Further research is also needed to explore these changes in adolescent and young adults who tend to engage in heavier drinking and in clinical populations. Other predictors of problematic alcohol use should also be investigated for their potential moderating influence, including consideration of mental health and environmental factors (e.g., family and peer influence, socioeconomic/geographic conditions, access to substances).

## Data Availability

The ABCD data used in this report came from https://doi.org/10.82525/jy7n-g441.

## Sources of Support

This project was supported by the National Institute on Alcohol Abuse and Alcoholism (NIAAA) under award number T32 AA013525 (VS). This project was also supported by the National Institute on Drug Abuse under award number F31 DA061618 (ED), K08 DA062011 (ALW), F32 DA064409 (RMS), and (DA050779, PI: Wade). The content is solely the responsibility of the authors and does not necessarily represent the official views of the NIH.

## Acknowledgements

Authors would like to thank the families who participate in the ABCD Study and the staff who collect the data. Data used in the preparation of this article were obtained from the <u>Adolescent Brain Cognitive Development™ (ABCD) Study</u>, held in the <u>NIH Brain</u> <u>Development Cohorts Data Sharing Platform</u>. This is a multisite, longitudinal study designed to recruit more than 10,000 children aged 9–10 and follow them over 10 years into early adulthood. The ABCD Study® is supported by the **National Institutes of Health** and additional federal partners under award numbers: U01DA041048, U01DA050989, U01DA051016, U01DA041022, U01DA051018, U01DA051037, U01DA050987, U01DA041174, U01DA041106, U01DA041117, U01DA041028, U01DA041134, U01DA050988, U01DA051039, U01DA041156, U01DA041025, U01DA041120, U01DA051038, U01DA041148, U01DA041093, U01DA041089, U24DA041123, U24DA041147. A full list of supporters is available at <u>Federal Partners – ABCD Study</u>. ABCD Consortium investigators designed and implemented the study and/or provided data but did not necessarily participate in the analysis or writing of this report. This manuscript reflects the views of the authors and may not reflect the opinions or views of the NIH or ABCD Consortium investigators. The ABCD data repository grows and changes over time. The ABCD data used in this report came from https://doi.org/10.82525/jy7n-g441.

## Notes

### Competing Interest Statement

The authors have declared no competing interest.

### Author Declarations

As a multi-site study with UC San Diego as the coordinating center, all sites rely on UCSD's IRB. UC San Diego's ABCD protocol number is #160091 ("The Adolescent Brain Cognitive Development (ABCD) Study: UC San Diego").Initial approval was issued Nov 01, 2020, with annual renewal sought.

